# The case series of 34 patients with COVID-19 diagnosed with HIV infection from Central and Eastern European Countries – Euroguidelines in Central and Eastern Europe Network Group data

**DOI:** 10.1101/2020.09.16.20191528

**Authors:** Kerstin Kase, Agata Skrzat-Klapaczyńska, Anna Vassilenko, Arjan Harxhi, Botond Lakatos, Gordana Dragović Lukić, David Jilich, Antonija Verhaz, Nina Yancheva, Florentina Dumitrescu, Raimonda Matulionyte, Andrzej Horban, Justyna D. Kowalska

## Abstract

**Background:** A novel coronavirus (SARS-CoV-2) causing coronavirus disease (COVID-19) was detected at the end of 2019 in China. There are many COVID-19 studies in progress however, little is known about the course of COVID-19 in people living with HIV (PLWH). The aim of our study was to describe epidemiology and clinical characteristics of PLWH diagnosed with COVID-19 reported form Central and Eastern European Countries.

**Methods:** On-line survey was sent to Euroguidelines in Central and Eastern Europe (ECEE) Network Group. Analysis included all confirmed COVID-19 cases between March 11 and June 26 2020 among PLWH in 12 countries: Albania, Belarus, Bosnia and Herzegovina, Bulgaria, Czech Republic, Estonia, Hungary, Lithuania, Poland, Romania, Russia, and Serbia.

**Results:** In total 34 cases were reported. The mean age of those patients was 42.7 years (IQR = 35.8-48.5) and most of the patients were male (70.6% vs 29.4%). The mean CD4+ T-cell count prior COVID-19 diagnosis was 558 cells/mm^3^ (IQR = 312-719) and HIV RNA viral load (VL) was undetectable in 18 of 34 (53%) cases, the data about most recent HIV RNA VL was not available in three cases (8,8%). Comorbidities were observed in 19 (55.9%) patients, mostly cardiovascular disease (27,8%), and in 10 (29.4%) patients had coinfection, mostly chronic hepatitis C (87.5%). The clinical course of COVID-19 was asymptomatic in 4 (12%) cases, mild disease without hospitalization was reported in 11 (32%) cases. Stable patients with respiratory and/or systemic symptoms have been documented in 14 (41%) cases; 5 (15%) patients were clinically unstable with respiratory failure. Full recovery was reported in 31 (91%) cases, two patients died. In one case the data was not available.

**Conclusion:** This study from 12 countries in Central and Eastern Europe region indicates no alarming signals of increased morbidity or mortality from COVID-19 among HIV-positive persons there is a need for further research.

## Introduction

The impact of COVID-19 infection on people living with HIV (PLWH) has not yet been fully understood. There is currently no scientific evidence suggesting an increased risk of acquiring the infection and a more severe course of the disease in HIV-infected patients, assuming that these patients have normal CD4 T-cell count and are successfully treated with antiretroviral therapy (cART). Generally, it has been clearly confirmed that older people and people with other comorbidities, including cardiovascular disease, type II diabetes and chronic pulmonary diseases are at higher risk of severe COVID-19 in the general population (1). HIV-infected individuals might be at higher risk of COVID-19 infection because significant proportion of them are over the age of 50, and the comorbidities such as cardiovascular disease and chronic lung disease are more common than in general population (2-4). Moreover, little is known about the course of the COVID-19 in PLWH. In our study, we describe epidemiology and clinical characteristics of HIV-positive patients diagnosed with COVID-19 reported form Central and Eastern European Countries.

## Material and methods

The Euroguidelines in Central and Eastern Europe Countries (ECEE) Network Group was established in February 2016 to review standard of care for HIV infection and viral hepatitis within the region. This retrospective analysis included all confirmed COVID-19 cases between March 11 and June 26 2020 among HIV-positive patients in 12 countries (Estonia, Czech Republic, Lithuania, Albania, Belarus, Romania, Serbia, Bosnia and Herzegovina, Poland, Russia, Hungary, Bulgaria). Anonymized data were collected through on-line survey (by the treating physicians). The dataset included age, sex, information about HIV infection (time since HIV diagnosis, baseline HIV RNA viral load (VL), most recent CD 4+ T-cell count prior COVID-19 infection, latest HIV VL prior COVID-19 infection, nadir CD 4+ T-cell count), information about cART regimen at the time of COVID-19 infection, information about comorbidities and coinfections, details about COVID-19 infection (course of the disease: asymptomatic, mild disease, symptomatic patients with respiratory, clinically unstable patients with respiratory failure, patients in critical condition and outcome).

### Ethical approvals

The study was approved by the Bioethical Committee of the Medical University of Warsaw (Nr AKBE/155/2020).

## Results

In total 34 COVID-19 cases among HIV-positive patients were reported. In 32 (94,1%) cases SARS-CoV-2 infection was confirmed by PCR examination (in 1 case IgG positive, in 1 case clinical course +antibody positivity IgM).

We documented 24 (70,6%) male patients, medium age within study cohort was 42.7 (IQR = 35.8–48.5). (Table 1).

Two (5,8%) patients were diagnosed with HIV infection because of COVID-19 diagnosis. Among 32 patients with known HIV status before COVID-19 the time since HIV diagnosis ranged between 0.1 to 22 years.

28 patients were on cART regimen at the time of COVID-19 diagnosis. Three (8,8%) patients were on PIs and 12 (35%) patients were on tenofovir (TDF or TAF) containing regimen. Six patients (17,6%) weren’t receiving any antiretroviral treatment at the time of SARS-CoV-2 infection.

The latest median of most recent CD4+ T-cell count prior COVID-19 infection was 558 cells/mm^3^ (IQR = 312–719). The most recent HIV RNA VL (before COVID-19) was < 50 copies/ml in 18 cases, the data about latest HIV VL was not available in three cases (8,8%). We observed comorbidities other than HIV infection in 19 (55,9%) patients. The most frequently seen comorbidities were cardiovascular disease in five (27,8%) cases, chronic lung disease or asthma in two (11,1%) cases, diabetes in two (11,1%) cases and obesity in two (11,1%) patients. Coinfection with hepatitis C has been documented in 10 cases (29,4%), coinfection with hepatitis B was reported in one case (2,9%) (Table 1).

Respiratory symptoms occurred in 27 (79,4%) cases while 26 (76,5%) patients presented general symptoms like fever, fatigue/malaise, muscle aches; only ten (29,4%) patients had gastrointestinal symptoms (diarrhea, nausea/vomiting, abdominal pain).

The most common symptoms of COVID-19 infection were cough in 24 (70,6%), fatigue/malaise in 24 (70,6%) and fever in 21 (61,8%) cases. We observed muscle aches in 17 (50%) cases, headache in 9 (26,5%) cases, loss of smell in 9 (26,5%) cases and loss of taste in 7 (20,6%) cases.

In our study asymptomatic course of COVID-19 was only seen in 4 (12%) cases, we reported mild disease without hospitalization in 11 (32%) cases. Stable patients with respiratory and/or systemic symptoms have been documented in 14 (41%) cases; we reported 5 (15%) clinically unstable patients with respiratory failure.

Generally, 22 (64,7%) patients were hospitalized. In three patients hospitalization on intensive unit care was needed, two of them were with detectable HIV RNA needed mechanical ventilation – one of these patients died. In our observation full recovery was reported in 31 (91%) cases, two patients died. In one case (2,9%) the data was not available (Table 1).

Table 1. HIV-related patients’ characteristics (attachment)

## Discussion

Our review of 34 cases of patients HIV-positive ensured that COVID-19 presents mostly as mild disease in majority of PLWH having full clinical recovery (91%). In the beginning of SARS-CoV-2 pandemic it could be assumed that in the PLWH group, a significant proportion of this population had an increased risk of developing a more severe form of COVID-19. This hypothesis was based of fact that PLWH population includes patients with a low CD 4 T cell count (< 200 cells / µl) and patients without cART, and therefore in this group are severely immunocompromised patients. In addition, according to the Centers for Disease Control and Prevention (CDC), the risk of immune suppression is unknown, but for other viral respiratory tract infections, in PLWH population the risk of infection is highest with low CD4 T-cell count and without cART (5). Published data has verified those assumptions. The comparable results to our study were reported by Harter et al. In this case series of 33 PLWH patients with COVID-19 infection it was observed that 91% of the patients recovered and 76% of patients have been classified as mild cases (6). Another study strongly supports the above data. Study cohort published by Gervasoni et al. consisting of 47 PLWH with COVID-19 infection 96% of patients has fully recovered (7). In the larger study on 51 group of patients HIV-infected diagnosed with COVID-19 in Madrid Vizcarra et al. found that only six (12 %) of studied patients were critically ill, among which two (4%) patients died (8). Other recent studies in small cohorts of patients also strongly support the above data (9).

In our study the most common symptoms of COVID-19 infection were cough in 24 (70,6%), fatigue/malaise in 24 (70,6%) and fever in 21 (61,8%) cases. In the largest description of COVID-19 in Europe, in prospective observational cohort study of 16 479 patients from 166 UK hospitals Docherty et al. presented cough (70%), fever (69%) and shortness of breath (65%) as the most common symptoms. Only four percent of cases reported no symptoms (10). In our review four (12%) patients were asymptomatic, which suggest even better clinical outcome of SARS-CoV-2 in PLWH than in general population.

International AIDS Society (IAS) and the World Health Organization (WHO) recommend PLWH to take the same precautions as the general population, as well as follow specific governmental recommendations. PLWH who know their serological status but have not yet received cART should have immediate antiretroviral treatment initiated (11). Unfortunately, in Central and Eastern European Countries access to cART is insufficient (12). We observed this fact in our study; only 82.6% of patients were on cART, but despite of that we didn’t reported increased morbidity and mortality from COVID-19 among PLWH.

We must admit, that the main limitation of our study is small group of patients. In this study from 12 countries in Central and Eastern Europe region we did not see any clear signals of increased morbidity or mortality caused by COVID-19 among HIV-positive persons. As the studies of SARS-CoV-2 among PLWH are performed in small study groups, there is a need for further research.

## Data Availability

All data is available if needed

## Notes

### Competing Interest Statement

The authors have declared no competing interest.

### Funding Statement

No external funding was received

